# Changes in ventilatory anaerobic threshold over time: How does the years of experience of the observer affect the results?

**DOI:** 10.64898/2025.12.05.25341675

**Authors:** Nanaho Minagawa, Kazuo Kondo, Hirotaka Nishijima, Takeo Ikota, Yusuke Mori

## Abstract

**Aims:** To determine the extent to which years of experience in anaerobic threshold (AT) assessment affects the reliability of AT determination at a regional general cardiovascular hospital under a strictly blinded protocol.

**Methods:** A controller was established to blind the data. The observers who determined AT were non-controller physical therapists working at the facility: three observers, with 4-17 years of experience, and one consultant physician with 30 years of experience. The data analyzed included 130 patients (260 data points) with cardiac disease who underwent a cardiopulmonary exercise test during the inpatient and maintenance phases. The observers determined AT twice from blinded and randomized data using the V-slope method.

**Results:** The inter-rater reliability (intra-class correlation coefficient [ICC]) of the AT assessment was good (0.87, range 0.87-0.94). The limits of agreement (LOA: two standard deviations of the difference between the two AT determinations, mL/min) ranged between 103.6-185.0. The overall change in AT over time was not significantly different between observers (66.6±121.4 mL/min; p<0.001); neither was the observers’ years of experience, ICC, or LOA. However, the magnitude of error in AT assessment in the inpatient and maintenance phases was significantly related to the error in change in AT over time (r=0.685-0.861, p<0.001).

**Conclusion:** AT was assessed under a blinded protocol by four observers with 4-30 years of experience at a cardiovascular hospital. The reliability of their AT determinations was high, and the change in AT over time readings did not differ significantly by the years of observer experience, ICC, or LOA.

## Introduction

The terms anaerobic threshold (AT), gas exchange threshold (GET), and ventilatory threshold VT), lactate threshold LT) are used in the context of a level of exercise intensity during an incremental exercise test where lactate levels increase in the blood compared to resting levels. Lactic acid (lactate + H^+^) is buffered by the bicarbonate ion (HCO3^-^) in the blood to form carbon dioxide (CO_2_). This excess CO_2_ is separate from the CO_2_ generated by mitochondrial energy metabolism. The point in the exercise test when this excess CO2 is generated is termed the AT (1).

AT has been used for decades as an index of exercise tolerance, but the current gold standard for determining the effect of exercise and cardiac disease prognosis is peak volume of oxygen (VO_2_) consumed during symptom-limited maximal exercise (2). However, peak VO_2_ can be affected by the effort put forth by the patient and may not be maximal effort. On the other hand, the most important feature of AT is that it is an index that is independent of the maximal effort by the patient, since it is detected well before maximal effort. If patient ID, clinical information, and type of intervention and its before/after status are fully blinded, the determined AT, even under observational studies, may be an objective measure of exercise tolerance (3,4). However, since detection and determination are subjective and depend on the observer, highly reliable measurement is required. Although methods for the detection of AT have already been established (1), data plots of actual expiratory parameters (e.g., VO_2_ vs. VCO_2_: the so-called slope graph) must be assessed for AT under noise caused by breathing. The baseline before the threshold is often not stable. In textbooks, typical examples relatively free of noise are chosen for the demonstration of basic concepts, but the raw plots in the field are truly “dangerous curves” (5). For this reason, some experience (familiarity) is considered necessary to determine AT.

The situation in which AT is most often used in routine cardiac rehabilitation is in determining changes in exercise tolerance over time. Regarding the assessment of AT change in response to an intervention (e.g., exercise therapy), the subjective influence (bias) of AT readings can first be addressed by blinded AT readings. The second issue is how much experience is needed regarding the reliability of the AT determination itself? Dolezal et al. (6) compared the AT assessments of two experienced instructors (E) and four novice respiratory fellows (N) who knew the basic concepts but had no experience in actual AT determination. The variability of their judgments (95% limits of agreement [LOA]: two standard deviations of the difference between the two determinations, mL/min) (7) widened from about 195 mL/min for the two E assessors to about four times more when AT assessments made by the four N assessors were included. Kaczmarek et al. (8) compared the AT assessments of two experienced physicians with several qualified medical assistants. The 95% LOA spread from 249 mL/min among physicians to just under twice that when including assistants. The conclusion is that simply understanding the basic principles of AT detection is not enough, because the reproducibility (reliability) of AT determination is greatly reduced when there is little experience in the field. Therefore, the objectives of this study were 1) to examine the relationship between differences in years of experience and inter- and intra-observer reliability of AT determination by four observers with different years of experience, and 2) to examine the extent to which differences in the reproducibility of AT assessments by individual observers affect the amount of change in AT over time. Also, the fact that this study was not conducted at a university hospital or other research institution, but with on-site physical therapists and physicians involved in cardiac rehabilitation at a general cardiovascular hospital site, distinguishes this study from previous studies.

## Methods

### 1. Study Population

#### a). Observer

In this study, one controller was assigned to randomize and blind the data to ensure complete blinding. The observers who assessed AT were either physical therapists who were working at the facility at the beginning of the study (three observers) or a consultant physician. All four observers were involved in cardiac rehabilitation in their daily practice, and the mean number of cardiopulmonary exercise tests (CPXs) performed during the data collection period was 163.5 per year: a mean value of 163.5/4 = approximately 40 tests per year per person while the controller was also on regular duty. The three observers had been performing CPXs for 17, 7, and 4 years. The consultant had approximately 30 years of experience in conducting CPXs and assessing AT. All observers were informed of the research methods at the beginning of the study, understood the purpose of the study, and agreed to participate in the study of their own free will. In addition, observers pledged in advance not to discuss AT determination with each other.

#### b). Participants

Of the 994 consecutively admitted patients with cardiovascular disease who underwent CPX at our facility between 2010 and 2020, clinical details of 130 cases with data available for both the inpatient and maintenance phases (n = 260) were extracted retrospectively. The number of cases required was calculated based on a previous study conducted at our hospital (9). Since the change in AT between the inpatient and maintenance phases was approximately 60 ± 200 mL/min, we aimed to employ this difference-value in our calculations and set the significance level at 5% and power at 80%. The number of required cases was set at 130, considering the presence of undetectable AT cases. The interval between the inpatient and maintenance phases was 3-6 months for all patients.

### 2. Steps in the implementation of blinded AT assessment

a). Breath-by-breath original patient data (one CSV file per patient) were transferred to Microsoft Excel software and converted to moving means of eight breaths. A V-slope graph was created using the VO_2_ and VCO_2_ values. The 260 patient data files were then randomized using a computer program and the file names were replaced with randomized numbers. In addition, personal information included in the file, such as IDs, names, height and weight, and measurement dates and times, were deleted. Two types of blinded files were needed to assess intra-observer reliability; therefore, randomization was performed twice, and two folders were prepared for analysis: one for the first reading and another for the second reading. Randomization was performed on the whole set of 260 data files (130 inpatient stage and 130 maintenance phase files combined).

b). Observers assessed AT by the V-slope method from the first analysis folder. For the V-slope method, a line parallel to the diagonal in the graph of VO_2_ and VCO_2_ (at a 45° angle to the x-axis) was used as the baseline, and the break point deviating from this baseline was judged as the AT. This modified V-slope method was devised by Sue et al. (10) and used in all case presentations in the textbook by Wasserman et al. (1), and also in the multicenter study by Myers et al. (11), the largest AT reliability study. Although this method is used in our facility for AT determination in daily practice, we re-reconfirmed it before distributing the data to the four observers for AT assessment. The AT values determined by the observers were entered into a separate Excel file and collected by the controller when all the first analyses had been completed. The same AT determination method was employed for the second folder of blinded case files. The AT determination period was set at 3 months for the first and second AT determinations, respectively.

c). The controller de-randomized and unblinded the data collected from each observer and performed the analysis.

### 3. Patient Background

Patient background including medications is shown in Table 1. The New York Heart Association (NYHA) functional class was not recorded in the medical database, but the physical therapist in charge of the cardiac rehabilitation program speculated that the patient cohort of this study was mostly NYHA class I or II. The data items obtained from the CPXs were analyzed for work rate, Borg scale score, HR, VO_2_, VCO_2_ and R, at the end of the study. Since the data collected in the study were examined retrospectively, and not all exercise tests were performed up to the symptomatic limit, the values at the end of the study were adopted (denoted as the final value). During the AT assessment, the actual values determined by each observer, as well as the number of cases for which a determination was possible (determination rate), and the indeterminate rate were noted. The difference in AT between the inpatient and maintenance periods was also calculated.

**Table 1.**
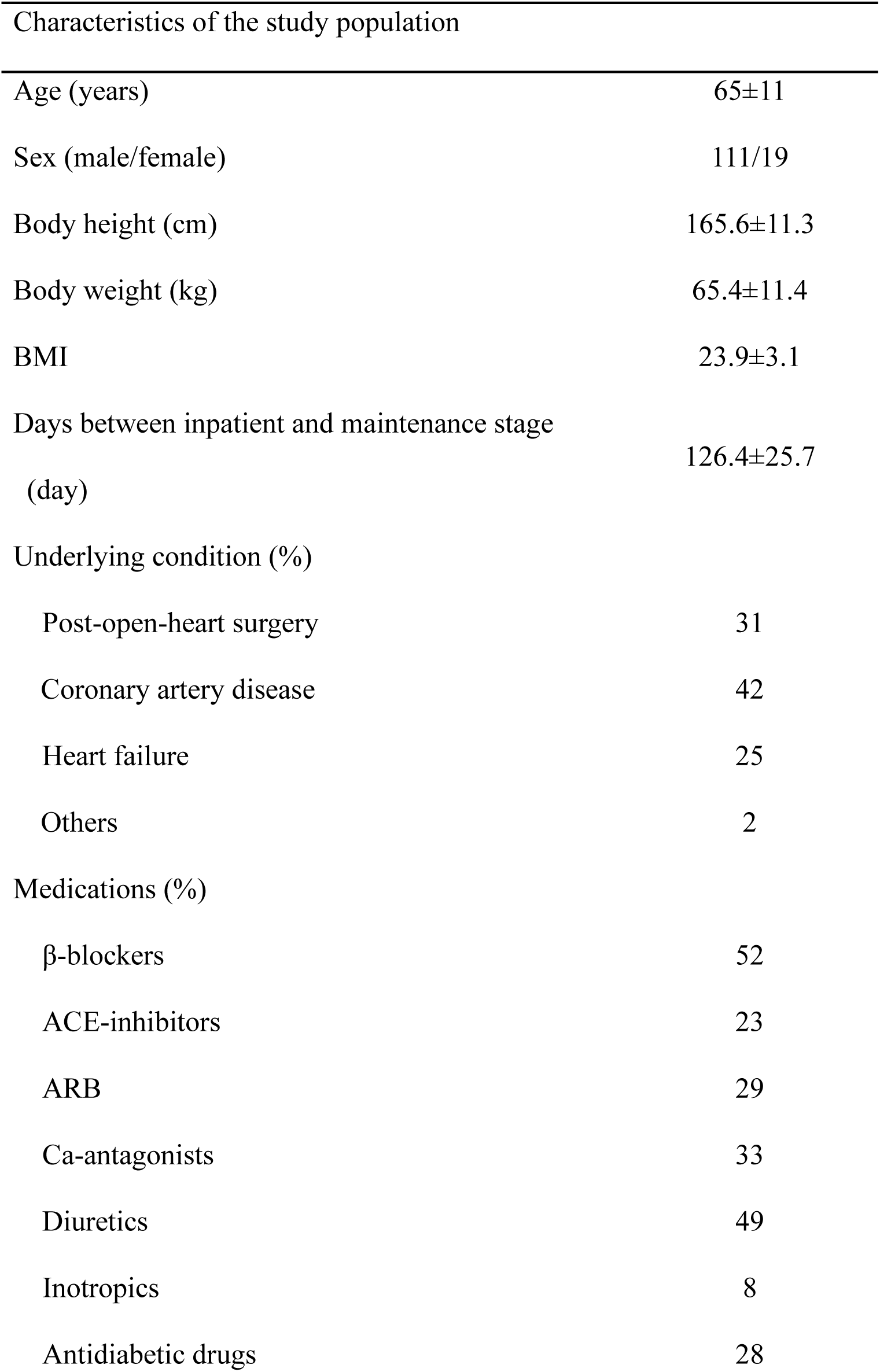
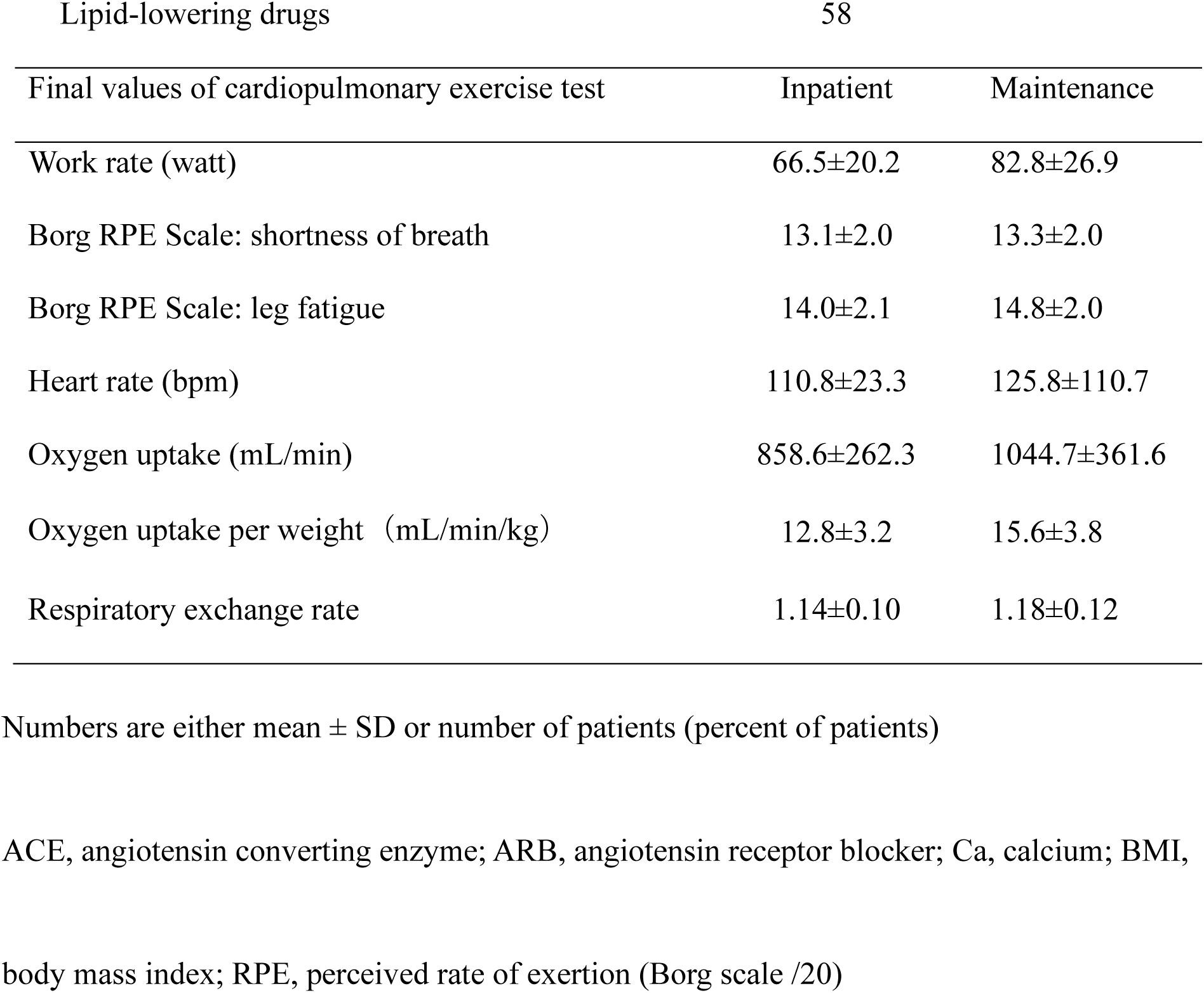
Characteristics of the study population and final values of the cardiopulmonary exercise test.

| Characteristics of the study population |  |
| --- | --- |
| Age (years) | 65±11 |
| Sex (male/female) | 111/19 |
| Body height (cm) | 165.6±11.3 |
| Body weight (kg) | 65.4±11.4 |
| BMI | 23.9±3.1 |
| Days between inpatient and maintenance stage<br>(day) | 126.4±25.7 |
| Underlying condition (%) |  |
| Post-open-heart surgery | 31 |
| Coronary artery disease | 42 |
| Heart failure | 25 |
| Others | 2 |
| Medications (%) |  |
| β-blockers | 52 |
| ACE-inhibitors | 23 |
| ARB | 29 |
| Ca-antagonists | 33 |
| Diuretics | 49 |
| Inotropics | 8 |
| Antidiabetic drugs | 28 |

| Lipid-lowering drugs |  | 58 |  |
| --- | --- | --- | --- |
| Final values of cardiopulmonary exercise test |  | Inpatient | Maintenance |
| Work rate (watt) |  | 66.5±20.2 | 82.8±26.9 |
| Borg RPE Scale: shortness of breath |  | 13.1±2.0 | 13.3±2.0 |
| Borg RPE Scale: leg fatigue |  | 14.0±2.1 | 14.8±2.0 |
| Heart rate (bpm) |  | 110.8±23.3 | 125.8±110.7 |
| Oxygen uptake (mL/min) |  | 858.6±262.3 | 1044.7±361.6 |
| Oxygen uptake per weight (mL/min/kg) |  | 12.8±3.2 | 15.6±3.8 |
| Respiratory exchange rate |  | 1.14±0.10 | 1.18±0.12 |

Numbers are either mean ± SD or number of patients (percent of patients)
ACE, angiotensin converting enzyme; ARB, angiotensin receptor blocker; Ca, calcium; BMI,
body mass index; RPE, perceived rate of exertion (Borg scale /20)

### 4. Statistical analysis

The AT determinations by the four observers were used to examine inter- and intra-observer reliability and margin of error in AT assessments. The effect of these parameters on the change in AT over time was also examined.

The intra-class correlation coefficient (ICC 2,1) was calculated for inter-observer reliability, and the intra-class correlation coefficient (ICC 1,1) and Bland-Altman limits of agreement (LOA) were calculated for intra-observer reliability (7). Correlation coefficients were calculated to see if LOAs differed by the observer’s years of experience. Changes in AT over time between the inpatient and maintenance phases were examined with a corresponding t-test for each observer and for the mean of the four observers. A repeated measures one-way analysis of variance (ANOVA) was calculated to determine the effect of inter-observer differences on the change in AT over time.

To examine the effect of the magnitude of error in AT determination during the inpatient and maintenance phases on the error in the change from inpatient to maintenance phases, the following terms were used. The inpatient stage was defined as Stage#1 (S1) and the maintenance phase as Stage#2 (S2), the first AT reading as Read#1 (R1) and the second, as Read#2 (R2). These abbreviations are summarized in Table 2. The first AT assessment in the inpatient stage was denoted as R1S1, the second AT determination in the inpatient period, as R2S1. Error (difference) in inpatient stage (dS1) = absolute value (R2S1 - R1S1), and the error (difference) in maintenance phase (dS2) = absolute value (R2S2 - R1S2). The sum of inpatient and maintenance phase errors (d total) was = dS1+dS2. The amount of change from inpatient to maintenance phase was expressed as cR1 = R1S2-R1S1, and cR2 = R2S2-R2S1) per two AT assessments. Based on the above, we examined the effect of the magnitude of error in AT assessment during the inpatient and maintenance phase (|dS1|+|dS2|) on the error in the amount of change from inpatient to maintenance phase (|cR2-cR1|).

**Table 2.** Abbreviations used in the estimation of the effect of reading error (difference) at Stage#1 and Stage#2 on the error of change between these two stages.

|  | Stage#1 | Stage#2 | Change |
| --- | --- | --- | --- |
| Read#1 | R1S1 | R1S2 | cR1(S2-S1) |
| Read#2 | R2S1 | R2S2 | cR2(S2-S1) |
| Difference | dS1(R2-R1) | dS2(R2-R1) |  |

Stage#1 and Stage#2 denote inpatient and maintenance stage, respectively.
Sum of error at Stage#1 and Stage#2 = $|dS1(R2-R1)| + |dS2(R2-R1)|$
Sum of error of change from stage#1 and Stage#2 = $|cR2(S2-S1) - cR1(S2-S1)|$
Note: cR1 and cR2, amount of change from the inpatient to maintenance phase, respectively;
dS1, error (difference) in the inpatient stage; dS2, error (difference) in the maintenance phase.

The consistency of the direction (improvement/worsening) of the two AT determinations from inpatient to maintenance phase was determined by the kappa coefficient. Statistical analyses were performed using R software version 4.1.2 (R Foundation for Statistical Computing, Vienna, Austria). The significance level was set at 5%. The ICC evaluation criteria followed Koo et al. (12).

### 5. Ethical Statement

Approval was obtained from the Ethics Committee of Hokko Memorial Hospital (Ethical Approval No. K-45) to conduct this study. This study was conducted in accordance with the Declaration of Helsinki and the Japanese Ethical Guidelines for Medical and Health Research involving Human Subjects. The data handled in this study were anonymized by the controller, who removed personal information and assigned random numbers so that individuals could not be identified. A table of correspondence between individuals and random numbers was kept by the controller. CPX was routinely performed after explaining the purpose and methods orally to the participants and confirming that sufficient understanding had been obtained.

## Results

### 1. Patient baseline characteristics and AT determination rate (Table 1 and 3)

The baseline characteristics of the 130 cases comprising our cohort are shown in Table 1. Most of the patients had ischemic heart disease, open heart surgery, or heart failure, and the mean duration of CPX from the inpatient to the maintenance phase was 126.4 ± 25.7 days. The basic parameters of CPX are shown in Table 1. The CPX protocol was performed at a ramp rate of 10 Watt/min in 94% of cases, but there were also cases performed at ramp rates of 5 Watt/min and 20 Watt/min. In all cases, the protocols for the inpatient and maintenance phases were identical. The AT determination rates ranged between 90.4 - 96.2% for each of the four observers for the 260 case files, and_all observers were able to determine AT in 225 cases,_or 86.5% of all cases (Table 3). Across inpatient and maintenance phases (130 cases), the four observers were able to determine AT in 83.1 - 92.3% of cases, and all observers were able to determine AT in 99 of the 130 cases, or 76.2% of all cases.

**Table 3.**
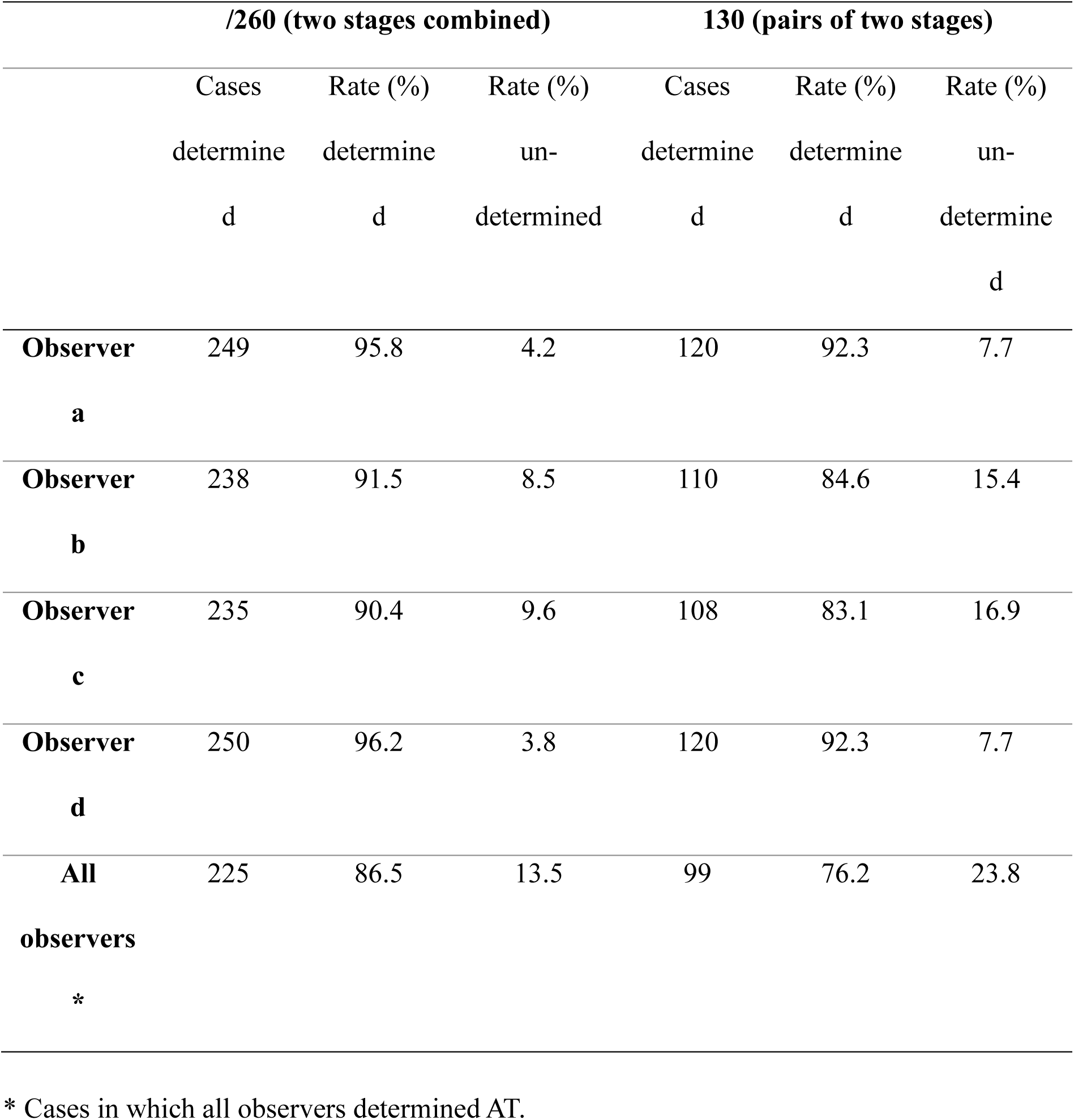
Determination of the rate of anaerobic threshold (AT)

### 2. Inter- and intra-observer reliability (Tables 4 and 5)

Inter-observer reliability was investigated in 99 cases (198 case files) and a determination was possible for all observers: (ICC 2,1) was 0.87 (95% confidence interval, 0.84-0.90). Intra-observer reliability, (ICC 1,1) ranged from 0.87-0.94 (95% confidence interval, 0.83-0.96) for the four observers, and LOA ranged from 103.6-185.0 mL/min.

**Table 4.** Inter-observer reliability.

| Inter-observer reliability | ICC (1,2) | 95% confidence interval |
| --- | --- | --- |
| n=198 | 0.87 | 0.84-0.90 |

| Inter-observer<br>reliability | n | ICC (2,1)<br>(95%CI) | Mean AT<br>(mL/min) | Difference<br>(mL/min) | LOA<br>(mL/min) | CV (%) |
| --- | --- | --- | --- | --- | --- | --- |
| Obs a vs. Obs b | 236 | 0.90 (0.84-0.93) | 616.2 | 33.2 | 157.8 | 22 |
| Obs a vs. Obs c | 233 | 0.83 (0.78-0.87) | 608.4 | 18.4 | 203.5 | 20 |
| Obs a vs. Obs d | 246 | 0.89 (0.86-0.92) | 609.6 | 22.3 | 166.0 | 22 |

Note: CV, coefficient of variation; ICC, intra-class coefficient; LOA, limits of agreement;
Obs, observer.

**Table 5.** Relationship between intra-observer reliability and change over time between inpatient and maintenance stages.

|  | ICC (1,1)<br>(95%CI) | Mean AT of<br>two stages<br>(mL/min) | Difference<br>(mL/min) | LOA<br>(mL/min<br>) | CV<br>(%) | Difference in<br>mean AT between<br>two stages<br>(mL/min) | p-<br>value |
| --- | --- | --- | --- | --- | --- | --- | --- |
| Obs a | 0.94<br>(0.93-0.96) | 596.0±187.<br>6 | 4.7±63.0 | 126.0 | 7 | 66.6 | <0.00<br>1 |
| Obs b | 0.93<br>(0.91-0.95) | 617.6±194.<br>8 | 35.4±51.8 | 103.6 | 6 | 55.4 | <0.00<br>1 |
| Obs c | 0.87<br>(0.83-0.89) | 622.2±178.<br>4 | 7.7±92.5 | 185.0 | 11 | 66.2 | <0.00<br>1 |
| Obs d | 0.94<br>(0.92-0.95) | 623.9±189.<br>9 | 4.5±67.5 | 135.0 | 8 | 74.3 | <0.00<br>1 |

Note: CV, coefficient of variation; ICC, intra-class coefficient; LOA, limits of agreement;
Obs, observer.

### 3. Changes in AT over time (Table 5)

The change in AT between the inpatient and maintenance phases (AT in maintenance phase minus AT in inpatient [mL/min]) showed a mean value of 66.6±121.4 mL/min (n=99) for all four observers, and the individual data for each observer also showed significant differences (p<0.001). The results of repeated measures of one-way ANOVA on this change in AT over time showed no significant differences between observers (p=0.74).

### 4. Relationship between intra-observer reliability and changes in AT over time (Figure 1 and Table 5)

Intra-observer reliability (ICC) ranged from 0.87-0.94 and LOA from 103.6-185.0 mL/min, but within this range there was no difference in AT over time. The years of experience of the four observers were not related to the ICC, LOA, or to the magnitude of changes over time (p=0.51). We assessed the direction of change in AT over time (improvement/worsening) between the first and second AT determinations. The opposite directions observed ranged from 11.2-17.9%, with kappa coefficients ranging from 0.592 - 0.760 (see Supplementary material online, Table S1). We also examined the effect of errors from two AT determinations during the inpatient and maintenance phases on the change in AT over time (difference between the inpatient and maintenance phases). The relationship between the total error of two readings of each stage (|dS1| + |dS2|) and the calculated difference in change over time (|cR2-cR1|) is shown in Figure 1. The total error and the error in change over time were significantly positively correlated (r=0.685 - 0.861 (n=4), each p<0.001). As shown in Figure 1, the error in change was equal to or less than the total error in AT determination during the inpatient and maintenance phases. The results were similar for other observers (see Supplementary material online, Figure S1). When the direction of the errors in the two AT determinations in the inpatient and maintenance phases was opposite, the difference, the change over time, was additive and maximum (i.e., equal to the total error in the inpatient and maintenance phases), and when the direction was the same, the difference was subtractive and less than the total error. Further explanation of this phenomenon is given in Supplementary material online, Figure S2 and S3 using actual data.

**Figure 1.**
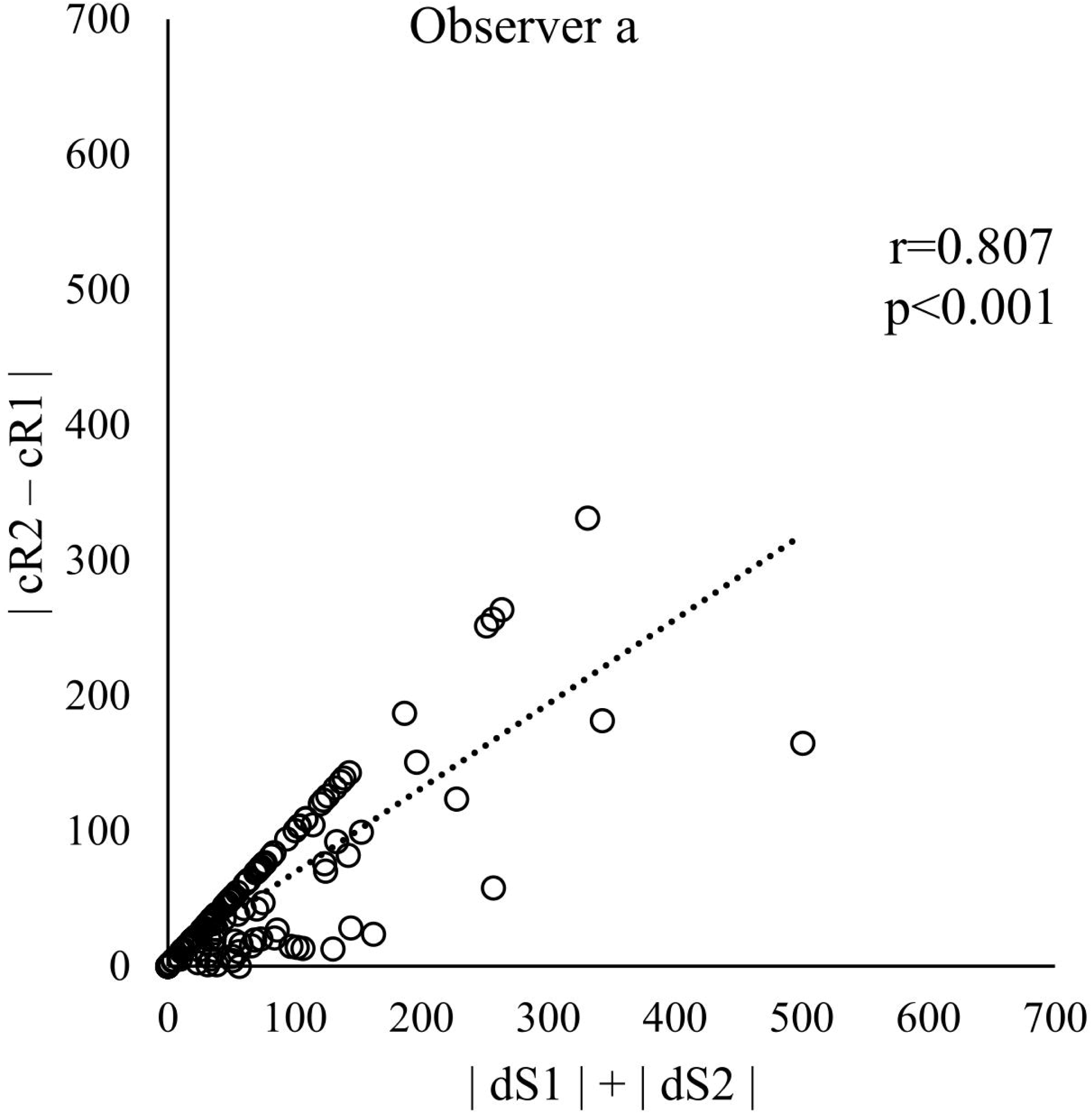
Effect of AT determination errors in the inpatient and maintenance phase on determination errors in the amount of change from inpatient to maintenance phase. Total AT error for inpatient and maintenance phase =|dS1|+|dS2|. Error in the amount of change from the inpatient to maintenance phase due to this error =|cR2-cR1|. Note: cR1 and cR2, amount of change from the inpatient to maintenance phase, respectively; dS1, error (difference) in the inpatient stage; dS2, error (difference) in the maintenance phase.

## Discussion

### 1) Reliability of AT determination and years of experience

With regard to the review of the literature on AT determinations, we included only those studies where the AT assessment was based on blinded patient data. Regarding the number of years of experience in assessing AT, it has been found to be necessary to obtain a certain degree of reproducibility (6,8), but the minimum number of years of experience is unclear. Conversely, the study by Abbot et al. (13) did not include newcomers to AT assessment contrary to two previous reports, but did not find significant differences in ICCs for years of AT or CPX experience (1-6 years or greater) and total number of cases (55-700 patients or more). The minimum number of years and number of cases of AT or CPX experience does not appear to be very high. Prior studies have incorporated the training of newcomers through materials and other means, but the effectiveness of such training also needs to be evaluated.

The three observers (physical therapists) in this study had varying years of experience (4, 7 and 17 years), yet these differences in years of experience did not affect the reliability of the AT assessments obtained in this study. The consultant physician with extensive experience, including publications in a small number of academic journals, had approximately 30 years of experience, but there was no difference in the reliability of his AT determination from the other three observers. In our study, there was no difference in the AT assessments of observers with 4 years or greater of experience, and no trend toward increased reliability with more years of experience beyond that.

### 2) Reproducibility of observers with experience

Regarding our review of the literature, the following conditions for inclusion were used: 1) it was clearly stated that the AT determination was blinded, and 2) ICC/LOA were considered. Dolezal et al. (6) performed three CPXs each on n=10 healthy participants, and the analysis showed a 95% LOA value of 195 mL/min (without ICC calculation). Kaczmarek et al. (8) performed CPX in 1,079 healthy participants and reported a 95% LOA = 249 mL/min and ICC=0.901. In a multicenter study, Myers et al. (11) performed 4 cycles of CPX in each of 428 patients with heart failure (NYHA III-IV, ejection fraction<=35%) and reported an intra-observer reliability (95% LOA) of 164 and 234 mL/min and inter-observer reproducibility (95% LOA) of 207 and 247 mL/min. Shimizu (14) reported an inter-observer reliability (ICC) of 0.93 (ergometer exercise, ramp protocol) in patients and participants (17 cardiac patients and 6 healthy participants, six times each with different exercise modalities and protocols). Dube et al. (15) reported that in healthy participants and patients with chronic obstructive pulmonary disease (COPD) (healthy n=23, COPD n=92), the V-slope method resulted in an inter-observer reliability (ICC) of 0.98 for healthy participants and ICC of 0.92 for patients with COPD in the Global Initiative for Obstructive Lung Disease-1 classification and a lower ICC of 0.78-0.35 for patients with COPD. Reproducibility appeared to be reduced in patients with advanced COPD.

There is no accepted or standard reference value for either ICC or LOA (16). However, the ICC value reported in the literature is predominantly 0.75 or higher, while for LOA this value ranges between 100-200 mL.

### 3) AT determination rate

In patients with cardiac disease, especially those with chronic heart failure, there are cases of exercise oscillatory ventilation, hyperventilation independent of AT, and indeterminate AT in patients with impaired exercise tolerance. Myers et al. (11) reported 16.4% of all patients with heart failure for whom neither of the two observers was able to determine AT. The indeterminate rate of AT in our present study was 4-10% of 260 case files, and 8-17% of 130 patients were indeterminate in either of the two AT determinations over time, with no significant difference among observers, and the results were similar to or less than those reported previously.

### 4) Error of AT at baseline and its relationship to change over time

The error of the calculated difference in AT between the inpatient and maintenance phases over time was found to be equal to or less than the total error of the inpatient and maintenance phases.

#### Limitations

In this study, AT determination was performed only by the V-slope method, so it was not possible to assess AT determination when other methods were used or when combinations of methods were used. Generally, the end-tidal VO_2_/VCO_2_, ventilatory equivalents of VO_2_ and VCO_2_ are also simultaneously used. Whether such a three-way decision method is more reliable or not needs to be examined. Also, consultation among observers has been reported. It is unproven whether this process produced better results. In addition, although the patients in the case files for our present study underwent CPX during the inpatient and maintenance phases, few of them received continuous outpatient rehabilitation during this period, and the majority of them received exercise prescriptions and unmonitored exercise therapy at the time of discharge from the hospital. Therefore, the results themselves do not reflect the therapeutic effect of a formal exercise program.

In this study, the inter- and intra-observer reliability of AT assessment in patients with cardiac disease who underwent blinded AT determination was high, and this was not affected by the years of observer experience (4-30 years) in assessing AT.

## Data Availability

All data produced in the present study are available upon reasonable request to the authors

## Acknowledgements

We thank all patients who underwent cardiopulmonary exercise tests during the study period, and the cardiac rehabilitation team of the hospital.

The original manuscript was written in Japanese. It was translated by using DeepL software.

The resulting manuscript was critically examined by the authors and was corrected where we thought there were errors and inappropriate rendering. We thank Editage for the professional English language editing of this manuscript.

## Authors’ contributions

NM, KK, HN contributed to the conception, or design of the work. NM, HN contributed to the acquisition, analysis or interpretation of data for the work. MN drafted the manuscript. NM, HN, KK, TI and YM revised the manuscript. All authors gave their final approval of the final version of the manuscript and agreed to be accountable for all aspects of this work, ensuring integrity and accuracy.

## Funding

None declared.

## Conflict of interests

The authors state that there are no conflicts of interest to declare.

## Data availability

The data underlying this article will be shared upon reasonable request made to the corresponding author.

---

**Supplemental Figure 1.**
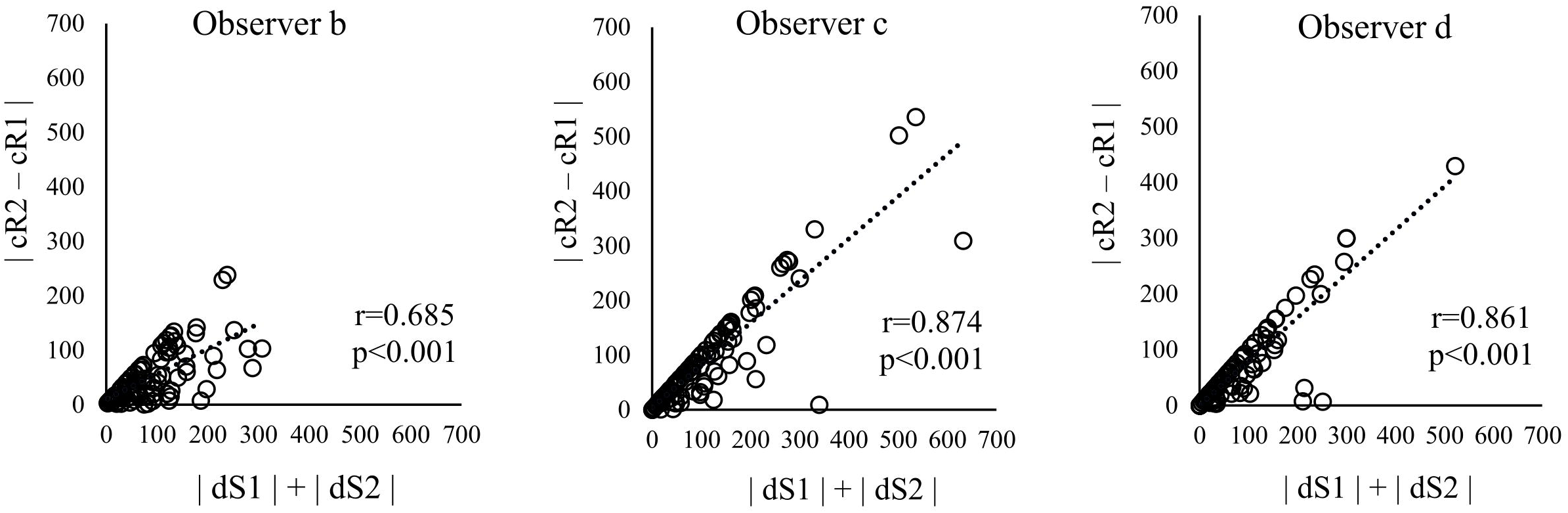
Effect of AT determination errors in the inpatient and maintenance phases on determination errors in the amount of change from the inpatient to maintenance phase (Observers b-d) Total AT error for inpatient and maintenance phase =|dS1|+|dS2|. Error in the amount of change from the inpatient to maintenance phase due to this error =|cR2-cR1|. Note: cR1 and cR2, amount of change from the inpatient to maintenance phase, respectively; dS1, error (difference) in the inpatient stage; dS2, error (difference) in the maintenance phase.

**Supplemental Figure 2.**
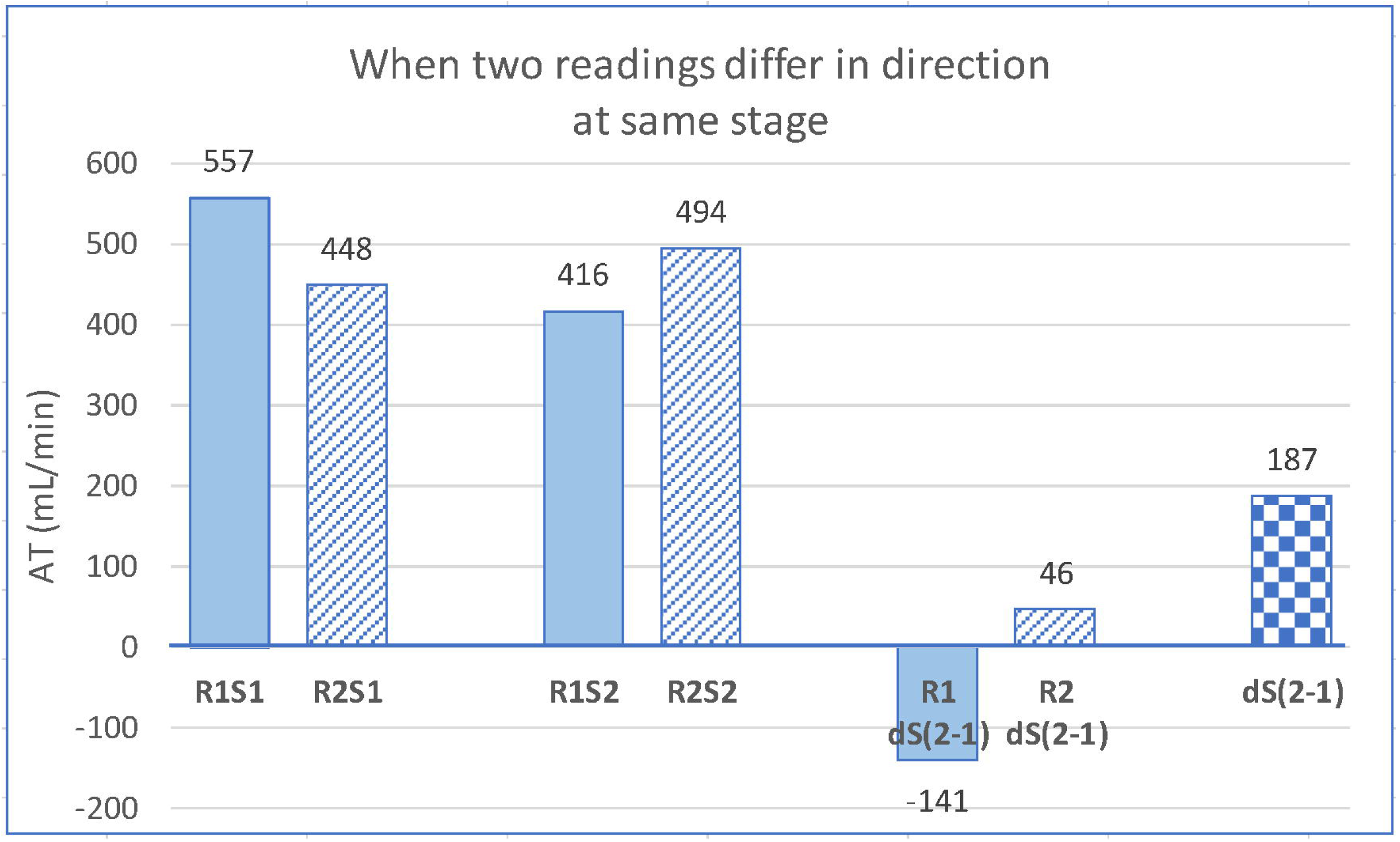
When two readings differ in direction at the same stage, the following equations apply. Total error of two stages(|dS1|+|dS2|) =|448-557|+|494-416| =|-109|+|78|=|187|=187 Change over the two stages (|cR2-cR1|) =|(494–448)-(416–557)|= |(46)-(−141)|= |46+141|=187 Note: cR1 and cR2, amount of change from the inpatient to maintenance phase, respectively; dS1, error (difference) in the inpatient stage; dS2, error (difference) in the maintenance phase.

**Supplemental Figure 3.**
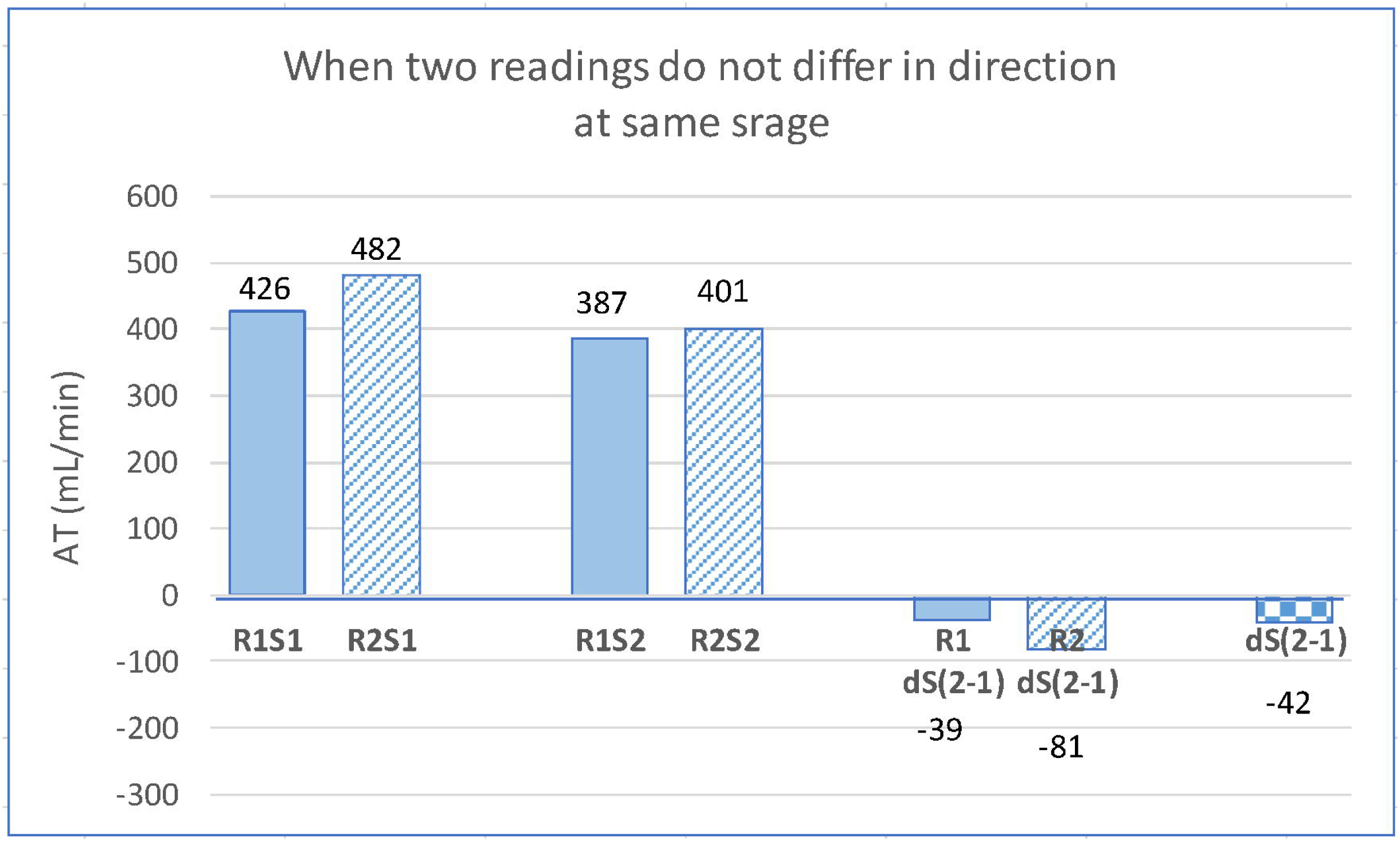
When two readings do not differ in direction at the same stage, the following equations apply. Total error of two stages(|dS1|+|dS2|) =|482-426|+|401-387| =|56|+|14|=|70|=70 Change over the two stages (|cR2-cR1|) =|(401–482)-(387–426)|= |(−81)-(−39)|= |-81+39)|=-42 Note: cR1 and cR2, amount of change from the inpatient to maintenance phase, respectively; dS1, error (difference) in the inpatient stage; dS2, error (difference) in the maintenance phase.

**Supplemental Table1.** Directional error in AT determination (LOA)

|  | LOA | Directional change between two stages |  | kappa |
| --- | --- | --- | --- | --- |
|  | (mL/min) | Agree (%) | Disagree (%) |  |
| Obs a | 159.7 | 88.2 | 11.8 | 0.759 |
| Obs b | 134.1 | 88.8 | 11.2 | 0.760 |
| Obs c | 257.3 | 82.1 | 17.9 | 0.592 |
| Obs d | 182.8 | 86.3 | 13.7 | 0.656 |

Note: AT, anaerobic threshold; LOA, limits of agreement; Obs, observer.

## Notes

### Competing Interest Statement

The authors have declared no competing interest.

### Funding Statement

This study did not receive any funding

### Author Declarations

Approval was obtained from the Ethics Committee of Hokko Memorial Hospital (Ethical Approval No. K-45) to conduct this study.

